# Association of Amygdala, Hippocampus, and Entorhinal cortex with thyroid function in older adults: Stratification’s value and relevance of bilateral volumetric analyses

**DOI:** 10.1101/2024.10.02.24314757

**Authors:** Asma Hallab, Alzheimer’s Disease Neuroimaging Initiative

**Affiliations:** Biologie Intégrative et Physiologie – Neurosciences Cellulaires et Intégrées. Faculté des Sciences et Ingénierie, Sorbonne Université, Paris, France; Pathologies du sommeil. Hôpital universitaire Pitié-Salpêtrière. Faculté de Médecine, Sorbonne Université, Paris, France; Charité - Universitätsmedizin Berlin, Corporate member of Freie Universität Berlin and Humboldt-Universität zu Berlin. Institute of Public Health. Berlin, Germany

**Keywords:** Ageing, Neurodegeneration, Psychoneuroendocrinology, Thyroid Stimulating Hormone, Magnetic Resonance Imaging, Cognitive Neurosciences

## Abstract

**Introduction:** Thyroid hormones modulate the brain structure during neurogenesis and impact cognition and emotions during the lifetime. It is, therefore, important to understand their association with relevant brain structures during the aging process.

**Methods:** A subset of 1348 older adults from the Alzheimer’s Disease Neuroimaging Initiative (ADNI) was included. Linear regression was used to study the association between serum thyroid stimulating hormone (TSH) and the Amygdala, Hippocampus, and Entorhinal cortex volumes. Sex and neurodegeneration-related stratifications and comparative bilateral volumetric analyses were performed.

**Results:** Females represented 667 (49%) of included cases, and 522 (38.72 %) were healthy controls (HC). A significant positive association was observed between TSH and total Hippocampus volume in mild cognitive impairment (MCI) (adj.*ß*=92 (23, 161), *p-value*=0.009), while a negative association in dementia participants remained statistically significant (*ß*=-177 (−295, −60), *p-value*=0.003 and adj.*ß*=-141 (−250, −32), *p-value*=0.012). There was a significant association between TSH and total Entorhinal cortex volume in the total study population (*ß*=44 (3.9, 85), *p-value*=0.032 and adj.*ß*=40 (5.1, 75), *p-value*=0.025). Stratification showed significant associations only in MCI (*ß*=80 (21, 138), *p-value*=0.007, and adj.*ß*=83 (27, 138), *p-value*=0.003), and males (adj.*ß*=54 (1.9, 106), *p-value*=0.042). Similar statistically significant associations were found only in the left Entorhinal cortex. The association between TSH and total Amygdala volume was positive in HC (*ß*=37 (1.6, 73), *p-value*=0.041) and negative in dementia participants (*ß*=-67 (−128, −6.4), *p-value*=0.030). None of those results remained statistically significant after adjusting the models. The bilateral volumetric analysis showed significant results only in the right Amygdala and dementia group.

**Conclusions:** Depending on the stratum and side of the volumetric analysis, significant associations were observed between TSH and Hippocampus, Amygdala, and Entorhinal cortex volumes. It is, therefore, crucial to consider the role of sex, neurodegeneration, and laterality when exploring the thyroid-brain interaction in older adults.

**Highlights:**

- Higher TSH levels are associated with lower Hippocampus volume on both sides in the dementia group.
- Lower TSH levels are associated with lower left Entorhinal cortex volume in the mild cognitive impairment and male strata.
- Higher TSH levels are associated with lower right Amygdala volume in the dementia group.

**Graphical abstract:** 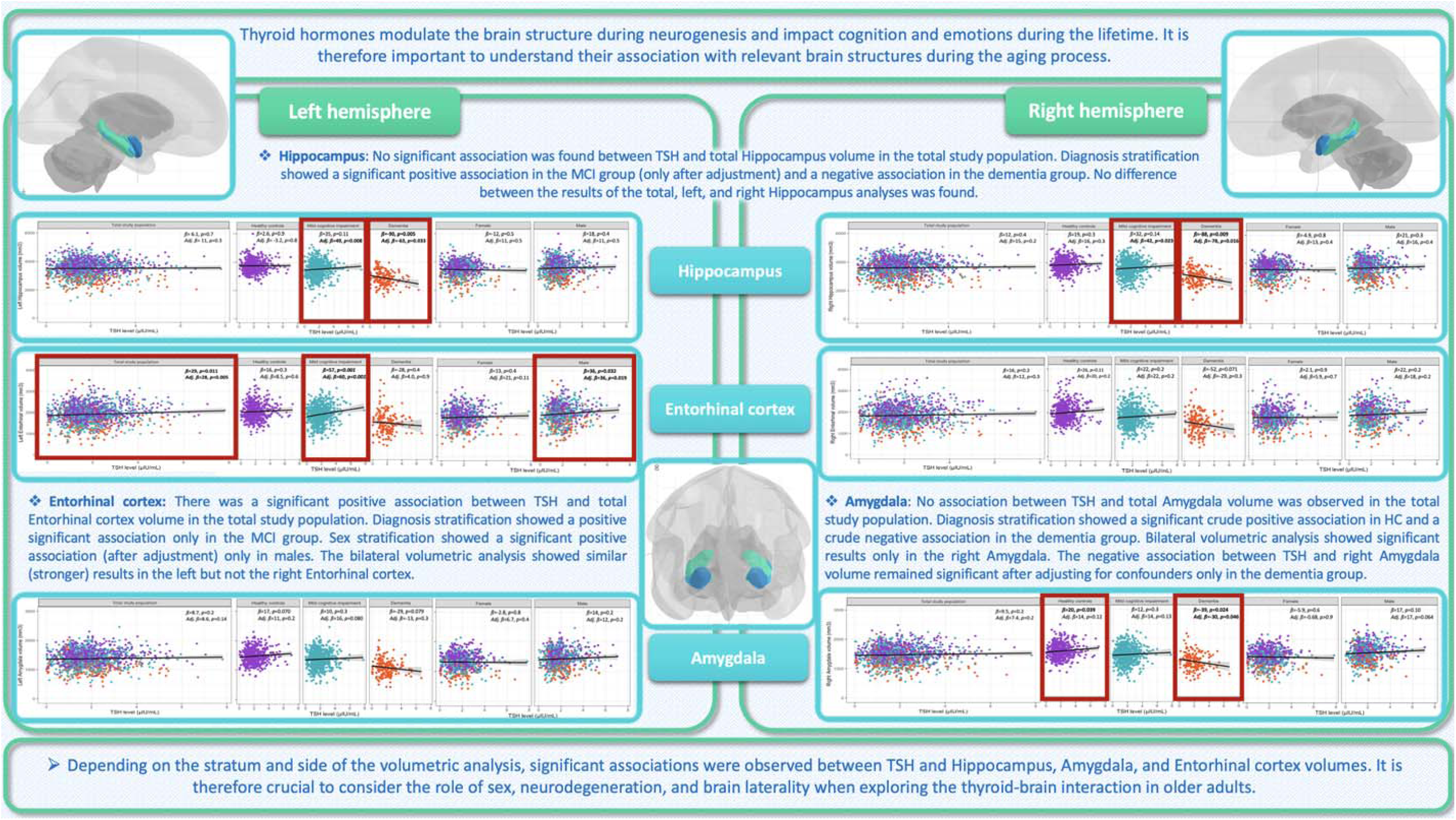

## 1. Introduction

The thyroid-brain association is a multifaceted interaction that, despite starting very early during gestational neurogenesis, remains relevant during adulthood and the aging process, making it of interest to all age spans and medical disciplines. (1)

Physiologically, thyroid function slowly declines during aging, resulting in normally increased thyroid stimulating hormone (TSH) levels in older adults. (2, 3) Similarly, aging is an independent risk factor for cognitive decline. (4) The association between thyroid function and cognitive decline in advanced ages has motivated several studies and raised various controversies, depending on studied populations, clinical definitions of thyroid dysfunction, and psychometric variables. (5, 6) While those studies put a particular focus on the association between hypothyroidism and cognitive decline, particularly in older adults, a smaller number of publications highlighted the association between lower TSH levels and cognitive impairment in the same age group. (7–9) Furthermore, fewer studies explored the association between thyroid function and anxiety, most of which assessed this interaction in the context of comorbid hypothyroidism and major depressive disorders in young adults. (10–12) Only one study on non-depressed older adults reported increased odds of anxiety in lower TSH ranges. (13)

It is, therefore, crucial to understand the underlying mechanisms defining the association between thyroid function and neuropsychiatric symptoms, particularly in older adults, a high-risk group of neurocognitive and neuropsychiatric adversities. Moreover, scientific evidence is in favor of a sex-modulated thyroid-brain interaction, and diverging effects might be observed between males and females. (7, 13, 14) Structural brain analysis is a relevant tool for understanding and quantifying volume loss and neurodegeneration. Limbic structures in the medial temporal lobe (MTL), mainly the Hippocampus and Entorhinal cortex, are particularly relevant centers for cognition and are early affected during the neurodegenerative process and dementia. (15, 16) Furthermore, the Amygdala, another limbic structure of the MTL, is associated with memory formation (16) and emotional processing and plays a pivotal role in experiencing and expressing particularly survival-relevant emotions such as fear. (17) It is also known that brain laterality has several implications for psychomotor and cognitive outcomes, and diverse factors, such as aging, might influence its structural and functional organization. (18)

Despite the largely discussed clinical evidence in favor of an interplay between thyroid and cognition across different age groups, very limited data is available on the association between thyroid function and brain structures in older adults. It is also unclear how sex and neurodegeneration might impact this association and whether brain function laterality might reflect structural discrepancies in the thyroid-brain interaction.

Therefore, the aim of the current study was: (A) to study the overall association between TSH and the total volumes of the Hippocampus, Entorhinal cortex, and Amygdala; (B) to evaluate the value of sex and neurodegeneration-dependent stratification in this association; and (C) to highlight the relevance of a bilateral volumetric approach.

## 2. Methods

The study was conceptualized and reported according to STrengthening the Reporting of OBservational studies in Epidemiology (STROBE) guidelines. (19)

### 2.1. Study population

The Alzheimer’s Disease Neuroimaging Initiative (ADNI) is a non-interventional longitudinal cohort initiated by the principal investigator Dr. Michael Weiner and funded by the National Institute on Aging (National Institutes of Health Grant U19 AG024904). The main objective of this cohort was to understand dementia and related risks. Healthy older adults, as well as those with cognitive impairment, were eligible. The recruitment of study participants took place in several centers across the United States of America and Canada. Neurocognitive, neuroimaging, and biological biomarkers were collected during recurrent study visits. Participants gave written consent. The study was performed according to the declaration of Helsinki, and ethical approvals were obtained from each local recruitment center’s IRB. Details and protocols can be found at https://adni.loni.usc.edu.

### 2.2. Thyroid function

TSH is a relevant biomarker of central thyroid function and was measured in fasting blood at baseline and reported in µIU/mL. Methods and data selection were detailed in previous publications. (7, 13) In summary, complete and accurate TSH measurements were individually screened, and only the first value was retained in case of duplicate measurements. Very low indetectable values (< 0.01 µIU/mL) were converted to 0.01 µIU/mL in three cases. Cases with TSH values equal to or higher than 10 µIU/mL (higher probability of overt hypothyroidism) were excluded from the analysis.

### 2.3. Neuroimaging and brain segmentations

ADNI participants underwent cerebral 1.5 or/and 3 Tesla magnetic resonance imaging (MRI) scans, depending on the study phase (ADNI 1 versus ADNI go, 2, and 3). Owing to variations in protocols and techniques used in each ADNI phase and recruitment center, details exceed the frame of this work, and protocols are published at https://adni.loni.usc.edu/data-samples/adni-data/neuroimaging/mri/. Several manuscripts described technical details of neuroimaging methods applied in different ADNI phases. (20, 21) For the current study, volumes of interest (VOIs) were selected based on a rigorous systematic review of the literature, and the volumetric analysis of T_1_-based segmentation was performed using FreeSurfer (https://surfer.nmr.mgh.harvard.edu). Longitudinal analyses (UCSFFSX51_11_08_19) for ADNI 1, go, and 2, and (UCSFFSX6_07_06_23) for ADNI 3 were used. Total Hippocampus volume was calculated as the sum of the volume of the left Hippocampus and right Hippocampus, total Entorhinal cortex volume as the sum of the volume of the left Entorhinal cortex and right Entorhinal cortex, and total Amygdala volume as the sum of the volume of the left Amygdala and right Amygdala. The total intracranial volume (ICV) was also reported and introduced to the multivariable regression models to adjust for anatomical variations. All volumes are measured in mm^3^.

### 2.4. Neuropsychological assessments

The cognitive status was mainly assessed using the total score of the Alzheimer’s Disease Assessment Scale – 13 items (ADAS_13_), in addition to total scores of Mini-Mental State Examination (MMSE), Clinical Dementia Rating scale – Sum of Boxes (CDR-SB), and Functional Activities Questionnaire (FAQ). Depression symptoms were assessed using the total score of the Geriatric Depression Scale (GDS). Anxiety symptoms were reported by study partners in the Neuropsychiatry Inventory Questionnaire (NPI-Q).

### 2.5. Inclusion criteria

Cases with missing TSH measurements (n=159), relevant VOIs (n=869 Hippocampus and n=4 Entorhinal cortex), main diagnosis at baseline (n=17), demographical data (n=4 missing age), ADAS_13_ total score (n=9), GDS total score (n=3), anxiety item in NPI-Q (n=10), biometric information for the BMI (n=3), or probably erroneous weight or height measurements (n=2), TSH ≥ 10 IU/mL (n=2) were removed allowing the inclusion of 1,348 cases.

### 2.6. Statistical analysis

RStudio version 2024.04.1-748 was applied for the analysis and visualization of the data. Values were presented as medians with interquartile ranges (IQR) or numbers with percentages (%) in continuous or count data, correspondingly. The associations between VOI volumes and TSH levels were assessed using linear regression analyses, where the VOI volume (mm^3^), as a continuous value, was the predicted variable, and continuous TSH levels (µIU/mL) as the predicting variable. A crude model was initially presented, and then the model was adjusted for relevant confounding factors:

- **Hippocampus and Entorhinal cortex-related adjusted models**: Age + sex + ADAS_13_ total score (points) + educational level (years) + APOE ε4 status (“None”, “one”, or “two alleles”) + cognition-related main diagnosis (“Healthy controls (HC)”, “Mild Cognitive Impairment (MCI)”, and “Dementia”) + ICV (mm^3^) + GDS total score (points).
- **Amygdala-related adjusted models** : Age + sex + ADAS_13_ total score (points) + educational level (years) + APOE ε4 status (“None”, “one”, or “two alleles”) + cognition-related main diagnosis (“HC”, MCI”, and “Dementia”) + ICV (mm^3^) + GDS total score (points) + Anxiety (binary).

The same strategy was followed after stratifying for sex (“Male” and “Female”) and cognition-related main diagnosis (“HC”, “MCI”, and “Dementia”). Adjusted models were adapted correspondingly after removing sex or main diagnosis when exploring the corresponding stratum. The total volume analyses were performed first; then, the analyses were performed with VOIs corresponding to the bilateral structures, each independently. Results were presented as regression coefficient (*ß*), 95% confidence interval (*CI*), and the *p*- value. The significance level of the two-sided *p*-value was set at 0.05. No correction for multiple testing was needed, as the current VOI analyses were based on peelable hypotheses driven by significant neurocognitive results in previously published studies from a larger subset of the same data, (7, 13) and no random search for statistical significance was followed during this work.

## 3. Results

### 3.1. Description of the study population

In addition to 522 (38.72 %) older HC, the study included 641 (47.55 %) with MCI and 185 (13.72 %) with dementia. Females represented 667 (49%) of the total population. The median serum TSH level was 1.74 µIU/mL (1.18 - 2.45). Detailed information on the study population and cognition-related subgroups are summarized in Table 1.

**Table 1:** Characteristics of the study population.

| *Characteristics* | *Overall*<br>*N = 1,348<sup>1</sup>* | *Healthy controls*<br>*N = 522<sup>1</sup>* | *Mild Cognitive Impairment*<br>*N = 641<sup>1</sup>* | *Dementia*<br>*N = 185<sup>1</sup>* |
| --- | --- | --- | --- | --- |
| **Age (years)** | 72 (67 - 77) | 70 (67 - 76) | 72 (66 - 77) | 75 (70 - 80) |
| **Sex** |  |  |  |  |
| *Female* | 667 (49%) | 304 (58%) | 288 (45%) | 75 (41%) |
| *Male* | 681 (51%) | 218 (42%) | 353 (55%) | 110 (59%) |
| **Racial profile** |  |  |  |  |
| *White* | 1,166 (86%) | 420 (80%) | 580 (90%) | 166 (90%) |
| *Black* | 114 (8.5%) | 67 (13%) | 37 (5.8%) | 10 (5.4%) |
| *Other* | 68 (5.0%) | 35 (6.7%) | 24 (3.7%) | 9 (4.9%) |
| **Marital status** |  |  |  |  |
| *Currently married* | 1,012 (75%) | 367 (70%) | 482 (75%) | 163 (88%) |
| *Currently not married or unknown* | 336 (25%) | 155 (30%) | 159 (25%) | 22 (12%) |
| **Housing situation** |  |  |  |  |
| *House or apartment* | 1,276 (95%) | 494 (95%) | 607 (95%) | 175 (95%) |
| *Retirement or nursing institution* | 49 (3.6%) | 20 (3.8%) | 22 (3.4%) | 7 (3.8%) |
| *Other* | 23 (1.7%) | 8 (1.5%) | 12 (1.9%) | 3 (1.6%) |
| **APOE ε4 status** |  |  |  |  |
| *0 allele* | 670 (56%) | 311 (69%) | 309 (53%) | 50 (29%) |
| *1 allele* | 422 (35%) | 125 (28%) | 215 (37%) | 82 (48%) |
| *2 alleles* | 113 (9.4%) | 16 (3.5%) | 59 (10%) | 38 (22%) |
| *Missing value* | 143 | 70 | 58 | 15 |
| **ADAS<sub>13</sub> total score (points)** | 12 (8 - 19) | 8 (5 - 12) | 14 (10 - 19) | 30 (24 - 36) |
| **MMSE total score (points)** | 29.00 (26.00 - 30.00) | 29.00 (28.00 - 30.00) | 28.00 (27.00 - 29.00) | 23.00 (21.00 - 25.00) |
| **FAQ total score (points)** | 0.0 (0.0 - 4.0) | 0.0 (0.0 - 0.0) | 1.0 (0.0 - 4.0) | 13.0 (8.0 - 18.3) |
| *Missing value* | 5 | 0 | 4 | 1 |
| **CDR-SB total score (points)** | 0.50 (0.00 - 2.00) | 0.00 (0.00 - 0.00) | 1.00 (0.50 - 2.00) | 4.50 (3.50 - 5.00) |
| **GDS total score (points)** | 1.00 (0.00 - 2.00) | 0.00 (0.00 - 1.00) | 1.00 (1.00 - 3.00) | 1.00 (1.00 - 3.00) |
| **Anxiety (informant-perceived)** | 166 (12%) | 16 (3.1%) | 99 (15%) | 51 (28%) |
| **BMI** | 26.6 (24.2 - 30.0) | 26.9 (24.3 - 30.4) | 26.6 (24.5 - 30.0) | 25.4 (22.8 - 28.3) |
| **TSH (μIU/mL)** | 1.74 (1.18 - 2.45) | 1.78 (1.22 - 2.50) | 1.68 (1.16 - 2.40) | 1.76 (1.10 - 2.49) |
| **Hippocampus total volume (mm<sup>3</sup>)** | 7,192 (6,395 - 7,856) | 7,599 (6,990 - 8,128) | 7,164 (6,383 - 7,783) | 5,835 (5,224 - 6,555) |
| **Entorhinal total volume (mm<sup>3</sup>)** | 3,761 (3,255 - 4,280) | 3,989 (3,594 - 4,456) | 3,720 (3,248 - 4,247) | 2,945 (2,437 - 3,480) |
| **Amygdala total volume (mm<sup>3</sup>)** | 2,851 (2,523 - 3,201) | 3,057 (2,726 - 3,300) | 2,824 (2,510 - 3,164) | 2,327 (1,954 - 2,594) |
| **ICV (mm<sup>3</sup>)** | 1,480,040 (1,377,036 - 1,592,153) | 1,459,980 (1,351,713 - 1,574,350) | 1,501,270 (1,398,670 - 1,602,620) | 1,470,461 (1,369,610 - 1,615,680) |

<sup>1</sup> Median (IQR); n (%)
**ADAS<sub>13</sub>:** Alzheimer's Disease Assessment scale – 13 items, **APOE:** Apolipoprotein, **BMI:** Body Mass Index, **CDR-SB:** Clinical Dementia Rating – Sum of Boxes, **FAQ:** Functional Activities Questionnaire, **GDS:** Geriatric Depression Scale, **ICV:** Intracranial volume, **MMSE:** Mini Mental Status Examination, **TSH:** Thyroid Stimulating Hormone.

### 3.2. Hippocampus and thyroid function

There was no association between serum TSH levels and total Hippocampus volume in the total study population (*ß_Total Hippocampus-Total population_*= 18 (−39, 74), *p-value*= 0.5 & adj. *ß_Total Hippocampus-Total population_* = 27 (−16, 70), *p-value*= 0.2).

After diagnosis stratification, significant associations were observed in cases with dementia (*ß_Total Hippocampus-Dementia_*= −177 (−295, −60), *p-value*= 0.003), but not in HC or those with MCI. After adjusting for confounders, a significant positive association was observed between serum TSH levels and total Hippocampus volume in MCI participants (adj. *ß_Total Hippocampus-MC_*=*_I_* 92 (23,161), *p-value*= 0.009). The negative association observed in participants with dementia remained statistically significant after adjustment for confounders (adj. *ß_Total Hippocampus-Dementia_* = −141 (−250, −32), *p-value*= 0.012).

The bilateral volumetric analysis showed similar results to those observed in the total Hippocampus volume in both left and right Hippocampi. Sex stratification did not show any significant difference between males and females. All details are visualized and summarized in Figure 1 and Supplementary Tables 1, 2, and 3.

**Figure 1:**
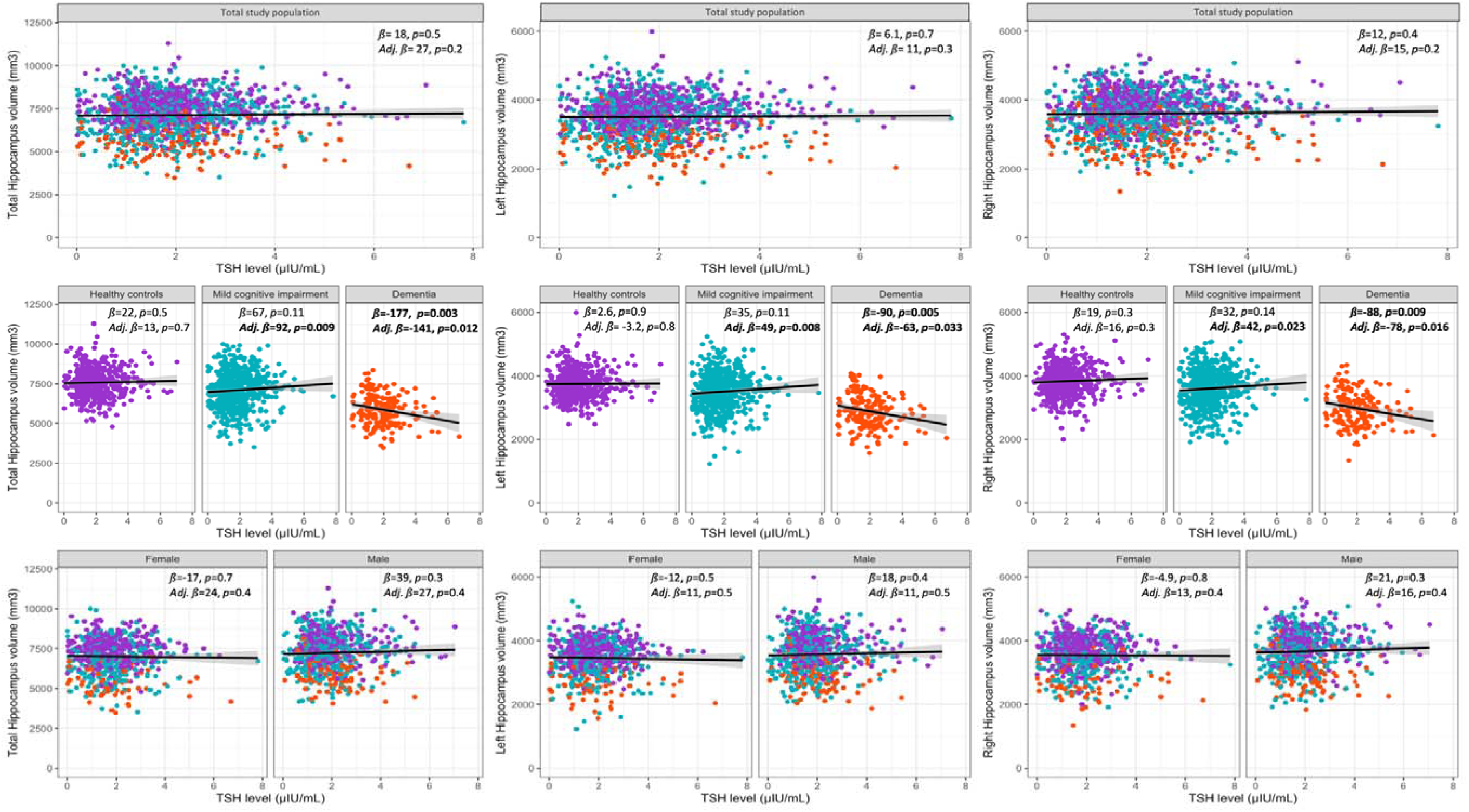
Crude and adjusted linear regression models on the association between TSH levels and Hippocampus volume, with sex and neurodegeneration-related stratifications and bilateral volumetric analyses.

### 3.3. Entorhinal cortex and thyroid function

There was a significant association between serum TSH levels and total Entorhinal cortex volume in the total study population (*ß_Total Entorhinal Cortex-Total population_* = 44 (3.9, 85), *p-value*= 0.032 & adj. *ß_Total Entorhinal Cortex-Total population_*= 40 (5.1, 75), *p-value*= 0.025).

The diagnosis stratification showed significant associations between thyroid function and total Entorhinal cortex volume only in MCI participants (*ß_Total Entorhinal Cortex-MC_*=*_I_* 80 (21, 138), *p-value*= 0.007 & adj. *ß_Total Entorhinal Cortex-MCI_* = 83 (27, 138), *p-value*= 0.003). After sex stratification, results remained statistically significant only in males (adj. *ß_Total Entorhinal Cortex-Males_*= 54 (1.9, 106), *p-value*= 0.042).

In the bilateral volumetric analyses, stronger statistically significant associations were found in the same strata of the left Entorhinal cortex but not the right one. Details are visualized in Figure 2 and Supplementary Tables 4, 5, and 6.

**Figure 2:**
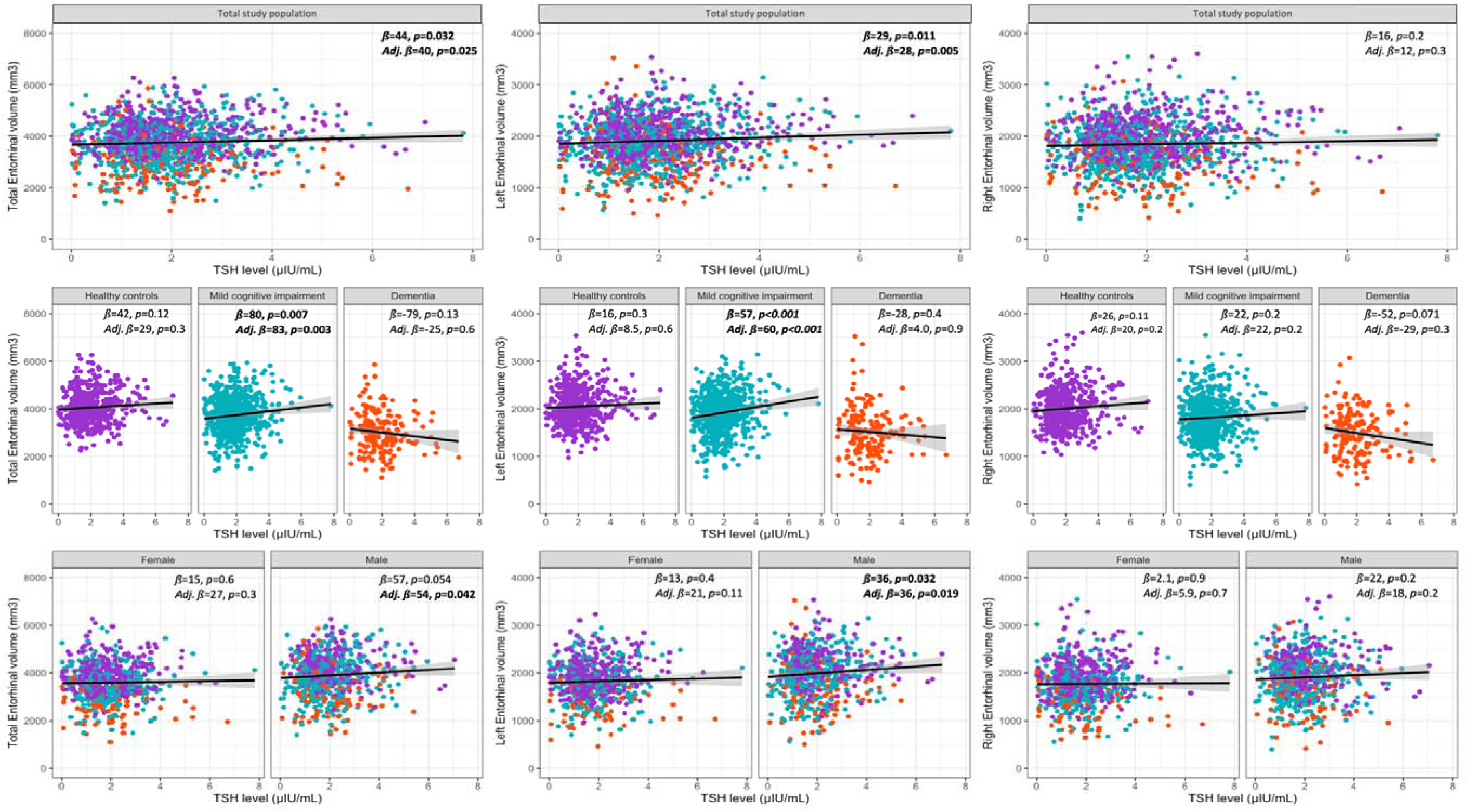
Crude and adjusted linear regression models on the association between TSH levels and Entorhinal cortex volume, with sex and neurodegeneration-related stratifications and bilateral volumetric analyses.

### 3.4. Amygdala and thyroid function

There was no significant association between serum TSH levels and the total Amygdala volume in the total study population.

While there was a positive association between thyroid function and total Amygdala volume in HC (*ß_Total Amygdala-HC_* = 37 (1.6, 73), *p-value*= 0.041), this association was negative in participants with dementia (*ß_Total_* _Amygdala-Dementia_= −67 (−128, −6.4), *p-value*= 0.030). None of those results remained statistically significant after adjusting the models for relevant confounders.

The bilateral volumetric analysis showed similar significant results in crude models only in the right Amygdala but not in the left one. The association remained statistically significant after adjusting for confounders in participants with dementia (*ß_Right Amygdala-Dementia_* = −39 (−72, - 5.2), *p-value*= 0.024 & adj. *ß_Right Amygdala-Dementia_* = −30 (−59, −0.56), *p-value*= 0.046). No significant associations were observed in sex strata.

The associations are visualized and summarized in Figure 3 and Supplementary Tables 7, 8, and 9.

**Figure 3:**
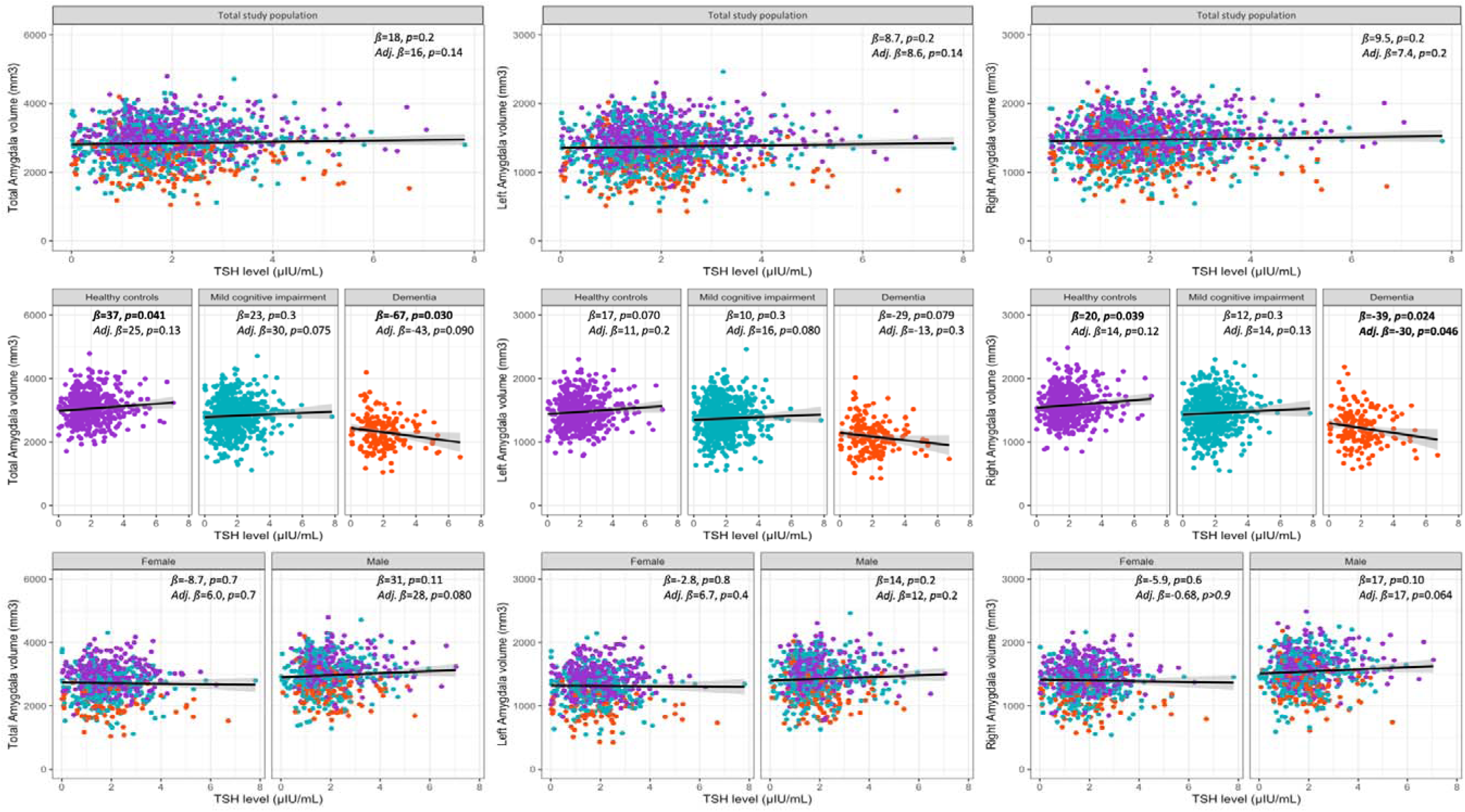
Crude and adjusted linear regression models on the association between TSH levels and Amygdala volume, with sex and neurodegeneration-related stratifications and bilateral volumetric analyses.

## 4. Discussion

The study explored the association between thyroid function and the volumes of three major limbic structures in older adults. The main outcome was the significant association between thyroid function and the Amygdala, Hippocampus, and Entorhinal cortex, depending on the studied sex- or neurodegeneration-related stratum. The bilateral VOI analyses highlighted the importance of exploring this association in left and right brain structures independently.

### 4.1. Thyroid and Hippocampus

A very limited number of published data studied the association between thyroid function and hippocampus volume, particularly in older adults. In a study including 62 premenopausal women with newly diagnosed Grave’s disease and hyperthyroidism, volumes of both hippocampi were significantly smaller compared to healthy controls. After treatment and correction of the hyperthyroidism, hippocampi volumes (right and left) increased significantly in size. There was also no correlation (Spearman) between hormone levels, particularly TSH, and hippocampi volumes. Patients with normal and pathological TSH levels did not show significant differences in hippocampi volumes during the follow-up. (22) In another study, adolescents with a history of congenital hypothyroidism showed smaller hippocampal volumes, particularly on the left side, compared to healthy controls. Furthermore, they neither showed a proportionated increase in hippocampal size with age compared to the control group nor a typical lateralization of cognitive functions. (23) On a longitudinal course, older adults with Alzheimer’s dementia but not healthy or MCI controls showed a significant association between lower TSH levels and annual hippocampal volume loss, particularly on the left side. (24)

In contrast with the longitudinal analysis, the association between TSH and bilateral hippocampus volumes was not statistically significant in the cross-sectional analysis at baseline. (24) In another study, newly diagnosed adults with subclinical or overt hypothyroidism presented with lower hippocampal subfield volumes, particularly on the right side, compared to HC. (25) Similarly, lower right hippocampal volumes were observed in adults with hypothyroidism compared to those without. (26) In a different population, antipsychotic-naïve patients with first-episode psychosis, high TSH levels were significantly correlated with lower total hippocampal volume at baseline. (27)

The results of published data are in favor of lower Hippocampus volume in both hyper- and hypothyroidism. Depending on the study design and the underlying population, both the left and/or right hippocampus might be affected. In contrast with previously published data, the majority of included cases in this study had TSH levels within the clinically normal range (IQR: 1.18 - 2.45 µIU/mL), and the two cases with eventual overt hypothyroidism were excluded. The three cases with very low TSH levels were converted to the lowest measurable value (0.01 µIU/mL) for the analysis.

In the current study, there were no relevant differences in the observed significance of the results between the right and left Hippocampi; significant associations were only observed in MCI participants after model adjustment and in participants with dementia, with and without model adjustment. The paradoxical difference between the observed positive association in the MCI group (higher TSH = higher Hippocampus volume) and the negative one in the dementia group (higher TSH = lower Hippocampus volume) remains unexplained.

Rodent studies have shown an association between thyroid hormones and gene expression in the Hippocampus, (28) their impact on neurogenesis and synaptic plasticity, (29, 30) as well as on defining white matter volume. (31)

### 4.2. Thyroid and Entorhinal cortex

Reviewing published data yielded no study exploring the association between thyroid function and the Entorhinal cortex in human subjects, particularly in older adults.

In the bilateral volumetric analysis of the Entorhinal cortex, the statistically significant results observed in the total study population remained significant only on the left side. Moreover, diagnosis stratification showed significant results only in the MCI group, while sex stratification showed significant results only in males.

The Entorhinal cortex, specifically the medial-lateral part, is the particular nest of grid cells coding in a complex computational architecture for spatial navigation and memory. (32, 33) In addition to those highly specific space cells, microglial integrity of the right Amygdala-Hippocampus-Entorhinal cortex was also associated with spatial learning in rodents. (34) The left Entorhinal cortex, in contrast, was found to be rather associated with verbal memory. (35)

Although the association between thyroid hormones and the Entorhinal cortex was rarely studied, a few rodent studies are available. One mechanism explaining the association of hypothyroidism with cognitive impairment is associated with the regulation of the Calcium-dependent calmodulin kinase II (CaMKII), a molecule present in the Entorhinal cortex and involved in learning and memory, by impairing the iodine-related phosphorylation. (36) The dysregulation of thyroid hormone signaling in the Entorhinal cortex is also linked to the risk of amyloid-ß toxicity observed in Alzheimer’s disease. (37) Furthermore, developmental hypothyroidism in rodents is associated with an impairment of the Entorhinal-Dentate gyrus neural pathway. (38)

### 4.3. Thyroid and cognition

The Hippocampus and Entorhinal cortex are critical brain structures that play a pivotal role in encoding and consolidating different areas of memory and learning. According to Braak staging, they are the first structures affected by tau and amyloid deposition and consequently by neurodegeneration during the aging process, specifically during Alzheimer’s dementia. (15)

A previous study including non-depressed healthy and MCI ADNI participants showed a significant association between lower TSH levels and worse cognitive performance in older males but not females. (7) Moreover, lower TSH levels predicted higher odds of being diagnosed with MCI at baseline. (7) Those previous observations go along with the current results, specifically, the significant positive associations between TSH levels and Entorhinal cortex volumes found in MCI and male strata.

### 4.4. Thyroid and amygdala

In the previously cited study on premenopausal women with Grave’s disease, amygdala volumes were significantly lower in affected women than in the control group (−10.4% for the left and −13.3% for the right Amygdala, both *p_t-test_*<0.001). After treatment, both increased significantly in size (+6.7% for the left and +11.1 for the right Amygdala, both *p_t- test_*<0.001). (22) Similarly to the Hippocampi volumes, no significant correlation was found between TSH and Amygdalae volumes. The only significant (negative) correlations in this population were observed between TSH receptor antibodies (TRAb) levels, on one side, and both Amygdala volumes and the right Hippocampus, on the other side. (22) In a different study, the administration of Levothyroxine (L-T_4_) to patients with bipolar disorder during a depressive episode was significantly associated with decreased metabolic activity in the right Amygdala and Hippocampus. (39)

### 4.5. Thyroid and anxiety

The amygdala plays a pivotal role in emotional processing, fear learning, empathy, and memory formation. (17, 40, 41) Dysfunction in Amygdalae is particularly associated with psychiatric disorders such as stress and affective disorders. (42–45)

A previous study including non-depressed healthy and cognitively impaired ADNI participants found a significant association between lower TSH levels and higher odds of informant-perceived anxiety. (13) After sex stratification, this association was only significant in older males but not females. (13) In the current study, cases with a total score of GDS higher than four points were included. The GDS was thus adjusted for in the regression models. Similarly, both cases with and without anxiety, as perceived and reported by their study partner, were eligible. In the regression models, where Amygdala volume was included as an outcome, anxiety was also adjusted for. It is, thus, important to mention that in the study on premenopausal women with Grave’s disease, no significant association between the level of anxiety and the volumes of the (right and left) Amygdalae was observed. (22)

The association between thyroid function and Amygdala volume was statistically significant only on the right side, particularly in older participants with dementia. The right Amygdala is known to be particularly involved in fear processing. (46) In young children, higher serum cortisol levels measured following a frustration task showed a significant association with lower right Amygdala volume, (47) which might be related to a chronic bilateral interplay between the right Amygdala and stress-related hormones. Furthermore, pathological connectivity patterns observed in the right Amygdala predicted suicidal risk in veterans. (48) In female patients, the severity of childhood abuse was negatively correlated with the right Amygdala volume, which was lower in those with posttraumatic stress disorder compared to HC. (49) Those studies are in favor of an association between Amygdala and stress.

Furthermore, in rodent studies, adult-onset hypothyroidism was associated with increased fear memory. (50) This association was explained by an enhanced Amygdala sensitivity to released mineralocorticoids and glucocorticoids mediated by an upregulation of corresponding receptor expression. (50) Moreover, thyroid hormones affect the expression of anxiety-related genes in the Amygdala. (51)

### 4.6. Strengths

The high number of included older participants, the sex and neurodegeneration stratification, and the bilateral volumetric analyses, are the main strengths of this study. This is the first study exploring the association between TSH and Entorhinal cortex and Amygdala volumes in older adults. The association related to both brain structures was of certain complexity since it showed dependence on the neurodegeneration state, sex, and the explored side of the brain. These particularities were detailed in the current analyses and opened the doors to further investigations aiming at a better understanding of the underlying mechanisms.

### 4.7. Limitations

The first limitation of the study is associated with the lack of free triiodothyronine (FT3) and free Thyroxine (FT4) measurements at baseline. While both hormones reflect peripheral thyroid function, the study’s main exposure was TSH as a biomarker for central thyroid function. The study included cases without extremely pathological TSH levels. Furthermore, TSH is negatively associated with FT_3_ and FT_4_ values and reflects therefore their variations, even within normal ranges. The second limitation is associated with the relatively high number of participants without data on VOI, which had to be excluded from the current analyses. Technical challenges and anatomical variations during neurodegeneration might explain this deficit. The third limitation is associated with the study design, seeing that cross-sectional analyses do not allow drawing a causal inference from the observed associations. Further longitudinal studies are needed.

## 5. Conclusions

The significant association between thyroid function and brain structures in older adults highlights the importance of the body-brain interaction during aging and the role of TSH in understanding the underlying process of cognition and emotions. The bilateral volumetric analysis showed lateralization in the association between thyroid function and different structures of MTL, as TSH levels predicted significant volume changes in the left Entorhinal cortex and right Amygdala. Furthermore, the degenerative remodeling observed in MCI and dementia might predispose to higher affinities between brain structures and TSH. Males and females might present further discrepancies in observed results, and the role of sex has to be taken into account when studying neurosciences in a hormonal context. Particularly in older adults, thyroid-brain interaction might be of complexity requiring multidisciplinary approaches, different strategies, and stratified analyses.

## Declarations

### Data availability

All data used in the manuscript is available at https://adni.loni.usc.edu.

### Author contributions

AH has full access to all of the data and takes responsibility for the integrity of the data and the accuracy of the analysis, visualization, drafting, and editing of the manuscript.

*Data used in preparation of this article were obtained from the Alzheimer’s Disease Neuroimaging Initiative (ADNI) database (adni.loni.usc.edu). As such, the investigators within the ADNI contributed to the design and implementation of ADNI and/or provided data but did not participate in analysis or writing of this report. A complete listing of ADNI investigators can be found at: http://adni.loni.usc.edu/wp-content/uploads/how_to_apply/ADNI_Acknowledgement_List.pdf.

## Acknowledgments

“Data collection and sharing for the Alzheimer’s Disease Neuroimaging Initiative (ADNI) is funded by the National Institute on Aging (National Institutes of Health Grant U19 AG024904). The grantee organization is the Northern California Institute for Research and Education. In the past, ADNI has also received funding from the National Institute of Biomedical Imaging and Bioengineering, the Canadian Institutes of Health Research, and private sector contributions through the Foundation for the National Institutes of Health (FNIH) including generous contributions from the following: AbbVie, Alzheimer’s Association; Alzheimer’s Drug Discovery Foundation; Araclon Biotech; BioClinica, Inc.; Biogen; Bristol-Myers Squibb Company; CereSpir, Inc.; Cogstate; Eisai Inc.; Elan Pharmaceuticals, Inc.; Eli Lilly and Company; EuroImmun; F. Hoffmann-La Roche Ltd and its affiliated company Genentech, Inc.; Fujirebio; GE Healthcare; IXICO Ltd.; Janssen Alzheimer Immunotherapy Research & Development, LLC.; Johnson & Johnson Pharmaceutical Research &Development LLC.; Lumosity; Lundbeck; Merck & Co., Inc.; Meso Scale Diagnostics, LLC.; NeuroRx Research; Neurotrack Technologies; Novartis Pharmaceuticals Corporation; Pfizer Inc.; Piramal Imaging; Servier; Takeda Pharmaceutical Company; and Transition Therapeutics.”

---

**Supplementary Table 1:** Crude and adjusted models of the regression analyses with <u>total Hippocampus volume</u> as the predicted variable and TSH as the predicting variable.

| Characteristics | Total study participants |  |  |  | Healthy controls |  |  |  | Mild cognitive impairment |  |  |  | Dementia |  |  |  | Females |  |  |  | Males |  |  |  |
| --- | --- | --- | --- | --- | --- | --- | --- | --- | --- | --- | --- | --- | --- | --- | --- | --- | --- | --- | --- | --- | --- | --- | --- | --- |
|  | N | Beta | 95% CI | p-value | N | Beta | 95% CI | p-value | N | Beta | 95% CI | p-value | N | Beta | 95% CI | p-value | N | Beta | 95% CI | p-value | N | Beta | 95% CI | p-value |
| **Crude models** |  |  |  |  |  |  |  |  |  |  |  |  |  |  |  |  |  |  |  |  |  |  |  |  |
| TSH (μIU/mL) | 1,348 | 18 | -39, 74 | 0.5 | 522 | 22 | -47, 90 | 0.5 | 641 | 67 | -16, 150 | 0.11 | 185 | -177 | -295, -60 | **0.003** | 667 | -17 | -95, 61 | 0.7 | 681 | 39 | -42, 120 | 0.3 |
| **Adjusted models** |  |  |  |  |  |  |  |  |  |  |  |  |  |  |  |  |  |  |  |  |  |  |  |  |
| TSH (μIU/mL) | 1,205 | 27 | -16, 70 | 0.2 | 452 | 13 | -46, 72 | 0.7 | 583 | 92 | 23, 161 | **0.009** | 170 | -141 | -250, -32 | **0.012** | 591 | 24 | -33, 81 | 0.4 | 614 | 27 | -37, 92 | 0.4 |
| Age (years) | 1,205 | -53 | -60, -46 | **<0.001** | 452 | -51 | -62, -40 | **<0.001** | 583 | -57 | -67, -47 | **<0.001** | 170 | -33 | -50, -16 | **<0.001** | 591 | -50 | -59, -41 | **<0.001** | 614 | -57 | -67, -46 | **<0.001** |
| Sex | 1,205 |  |  |  | 452 |  |  |  | 583 |  |  |  | 170 |  |  |  | 591 |  |  |  | 614 |  |  |  |
| Female |  | — | — |  |  | — | — |  |  | — | — |  |  | — | — |  |  | — | — |  |  | — | — |  |
| Male |  | 274 | 161, 387 | **<0.001** |  | 60 | -116, 235 | 0.5 |  | 355 | 183, 527 | **<0.001** |  | 113 | -195, 421 | 0.5 |  |  |  |  |  |  |  |  |
| Racial profile | 1,205 |  |  |  | 452 |  |  |  | 583 |  |  |  | 170 |  |  |  | 591 |  |  |  | 614 |  |  |  |
| White |  | — | — |  |  | — | — |  |  | — | — |  |  | — | — |  |  | — | — |  |  | — | — |  |
| Black |  | -102 | -321, 117 | 0.4 |  | -93 | -350, 164 | 0.5 |  | 24 | -354, 402 | 0.9 |  | -324 | -1,112, 464 | 0.4 |  | 45 | -203, 293 | 0.7 |  | -274 | -679, 132 | 0.2 |
| Other |  | 120 | -113, 353 | 0.3 |  | 131 | -183, 446 | 0.4 |  | 111 | -266, 487 | 0.6 |  | 49 | -564, 662 | 0.9 |  | 171 | -113, 456 | 0.2 |  | 100 | -285, 485 | 0.6 |
| Education (years) | 1,205 | -13 | -32, 6.4 | 0.2 | 452 | -42 | -70, -14 | **0.003** | 583 | -5.7 | -34, 23 | 0.7 | 170 | 45 | -9.8, 99 | 0.11 | 591 | -29 | -55, -2.5 | **0.032** | 614 | -2.5 | -31, 26 | 0.9 |
| Diagnosis | 1,205 |  |  |  |  |  |  |  |  |  |  |  |  |  |  |  | 591 |  |  |  | 614 |  |  |  |
| HC |  | — | — |  |  | — | — |  |  | — | — |  |  | — | — |  |  | — | — |  |  | — | — |  |
| MCI |  | -288 | -407, -169 | **<0.001** |  |  |  |  |  |  |  |  |  |  |  |  |  | -260 | -420, -100 | **0.002** |  | -347 | -525, -169 | **<0.001** |
| Dementia |  | -681 | -909, -453 | **<0.001** |  |  |  |  |  |  |  |  |  |  |  |  |  | -449 | -779, -120 | **0.008** |  | -872 | -1,191, -554 | **<0.001** |
| APOE ε4 (number) | 1,205 | -93 | -168, -18 | **0.015** | 452 | -74 | -193, 46 | 0.2 | 583 | -64 | -174, 46 | 0.3 | 170 | 4.0 | -187, 195 | >0.9 | 591 | -75 | -176, 26 | 0.14 | 614 | -91 | -202, 20 | 0.11 |
| ADAS<sub>13</sub> (points) | 1,205 | -41 | -49, -34 | **<0.001** | 452 | -22 | -38, -5.7 | **0.008** | 583 | -57 | -69, -46 | **<0.001** | 170 | -17 | -33, -1.7 | **0.030** | 591 | -46 | -56, -35 | **<0.001** | 614 | -38 | -49, -26 | **<0.001** |
| GDS (points) | 1,205 | -3.7 | -39, 31 | 0.8 | 452 | -38 | -101, 24 | 0.2 | 583 | -3.5 | -53, 45 | 0.9 | 170 | 83 | -3.2, 169 | 0.059 | 591 | -21 | -67, 26 | 0.4 | 614 | 15 | -38, 68 | 0.6 |
| BMI | 1,205 | 18 | 8.2, 27 | **<0.001** | 452 | 15 | 1.9, 28 | **0.025** | 583 | 24 | 9.5, 38 | **0.001** | 170 | -6.7 | -34, 21 | 0.6 | 591 | 20 | 8.6, 30 | **<0.001** | 614 | 13 | -3.8, 30 | 0.13 |
| ICV (mm<sup>3</sup>) | 1,205 | 0.00 | 0.00, 0.00 | **<0.001** | 452 | 0.00 | 0.00, 0.00 | **<0.001** | 583 | 0.00 | 0.00, 0.00 | **<0.001** | 170 | 0.00 | 0.00, 0.00 | **<0.001** | 591 | 0.00 | 0.00, 0.00 | **<0.001** | 614 | 0.00 | 0.00, 0.00 | **<0.001** |

ADAS<sub>13</sub>: Alzheimer's Disease Assessment Scale – 13 items, APOE: Apolipoprotein E, BMI: Body Mass Index, CI: Confidence Interval, GDS: Geriatric Depression Scale, HC: Healthy Controls, ICV: Intracranial volume, MCI: Mild Cognitive Impairment, TSH: Thyroid Stimulating Hormone.

**Supplementary Table 2:**
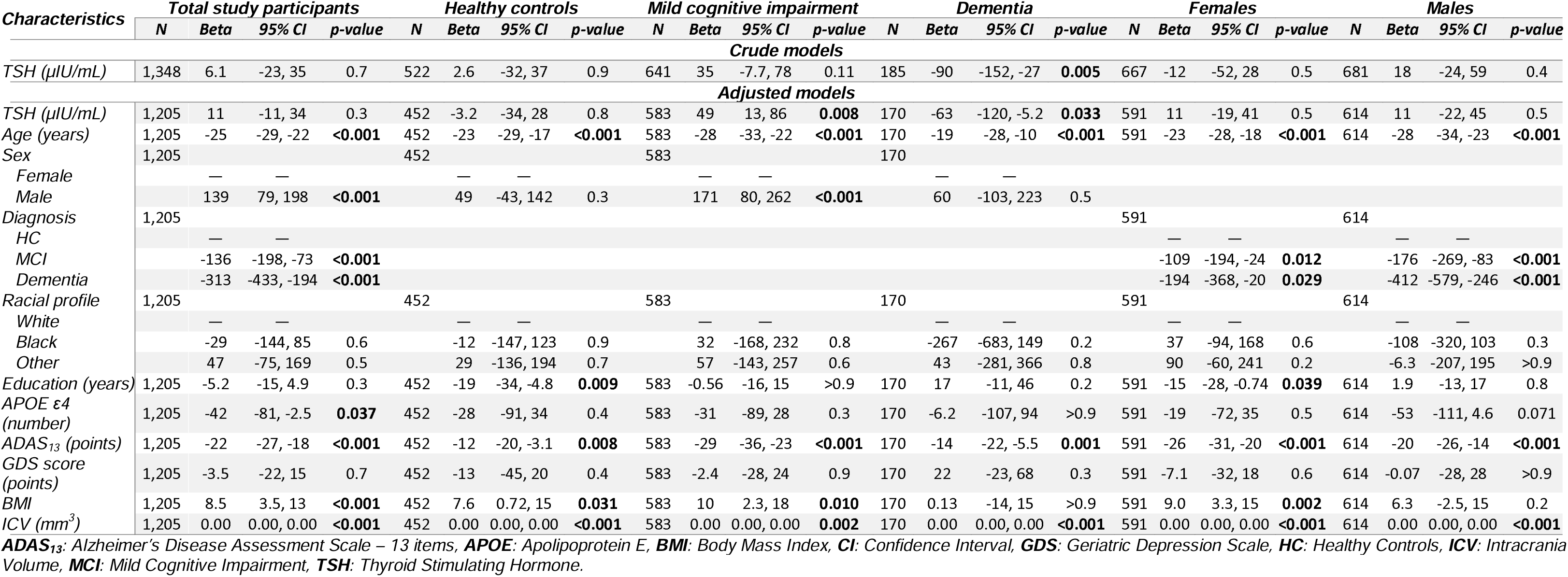
Crude and adjusted models of the regression analyses with <u>left Hippocampus volume</u> as the predicted variable and TSH as the predicting variable.

**Supplementary Table 3:**
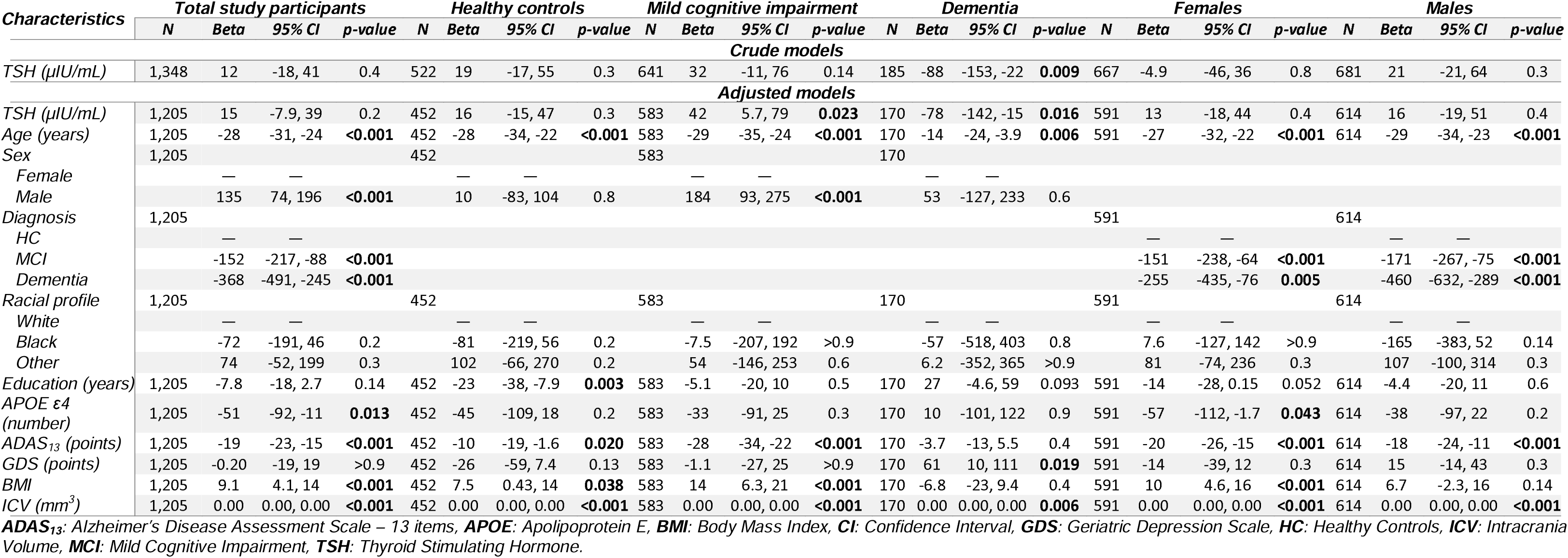
Crude and adjusted models of the regression analyses with <u>right Hippocampus volume</u> as the predicted variable and TSH as the predicting variable.

| Characteristics | Total study participants |  |  |  | Healthy controls |  |  |  | Mild cognitive impairment |  |  |  | Dementia |  |  |  | Females |  |  |  | Males |  |  |  |
| --- | --- | --- | --- | --- | --- | --- | --- | --- | --- | --- | --- | --- | --- | --- | --- | --- | --- | --- | --- | --- | --- | --- | --- | --- |
|  | N | Beta | 95% CI | p-value | N | Beta | 95% CI | p-value | N | Beta | 95% CI | p-value | N | Beta | 95% CI | p-value | N | Beta | 95% CI | p-value | N | Beta | 95% CI | p-value |
| **Crude models** |  |  |  |  |  |  |  |  |  |  |  |  |  |  |  |  |  |  |  |  |  |  |  |  |
| TSH (µIU/mL) | 1,348 | 12 | -18, 41 | 0.4 | 522 | 19 | -17, 55 | 0.3 | 641 | 32 | -11, 76 | 0.14 | 185 | -88 | -153, -22 | **0.009** | 667 | -4.9 | -46, 36 | 0.8 | 681 | 21 | -21, 64 | 0.3 |
| **Adjusted models** |  |  |  |  |  |  |  |  |  |  |  |  |  |  |  |  |  |  |  |  |  |  |  |  |
| TSH (µIU/mL) | 1,205 | 15 | -7.9, 39 | 0.2 | 452 | 16 | -15, 47 | 0.3 | 583 | 42 | 5.7, 79 | **0.023** | 170 | -78 | -142, -15 | **0.016** | 591 | 13 | -18, 44 | 0.4 | 614 | 16 | -19, 51 | 0.4 |
| Age (years) | 1,205 | -28 | -31, -24 | **<0.001** | 452 | -28 | -34, -22 | **<0.001** | 583 | -29 | -35, -24 | **<0.001** | 170 | -14 | -24, -3.9 | **0.006** | 591 | -27 | -32, -22 | **<0.001** | 614 | -29 | -34, -23 | **<0.001** |
| Sex | 1,205 |  |  |  | 452 |  |  |  | 583 |  |  |  | 170 |  |  |  | 591 |  |  |  | 614 |  |  |  |
| Female |  | — | — |  |  | — | — |  |  | — | — |  |  | — | — |  |  |  |  |  |  |  |  |  |
| Male |  | 135 | 74, 196 | **<0.001** |  | 10 | -83, 104 | 0.8 |  | 184 | 93, 275 | **<0.001** |  | 53 | -127, 233 | 0.6 |  |  |  |  |  |  |  |  |
| Diagnosis | 1,205 |  |  |  |  |  |  |  |  |  |  |  |  |  |  |  | 591 |  |  |  | 614 |  |  |  |
| HC |  | — | — |  |  |  |  |  |  |  |  |  |  |  |  |  |  | — | — |  |  | — | — |  |
| MCI |  | -152 | -217, -88 | **<0.001** |  |  |  |  |  |  |  |  |  |  |  |  |  | -151 | -238, -64 | **<0.001** |  | -171 | -267, -75 | **<0.001** |
| Dementia |  | -368 | -491, -245 | **<0.001** |  |  |  |  |  |  |  |  |  |  |  |  |  | -255 | -435, -76 | **0.005** |  | -460 | -632, -289 | **<0.001** |
| Racial profile | 1,205 |  |  |  | 452 |  |  |  | 583 |  |  |  | 170 |  |  |  | 591 |  |  |  | 614 |  |  |  |
| White |  | — | — |  |  | — | — |  |  | — | — |  |  | — | — |  |  | — | — |  |  | — | — |  |
| Black |  | -72 | -191, 46 | 0.2 |  | -81 | -219, 56 | 0.2 |  | -7.5 | -207, 192 | >0.9 |  | -57 | -518, 403 | 0.8 |  | 7.6 | -127, 142 | >0.9 |  | -165 | -383, 52 | 0.14 |
| Other |  | 74 | -52, 199 | 0.3 |  | 102 | -66, 270 | 0.2 |  | 54 | -146, 253 | 0.6 |  | 6.2 | -352, 365 | >0.9 |  | 81 | -74, 236 | 0.3 |  | 107 | -100, 314 | 0.3 |
| Education (years) | 1,205 | -7.8 | -18, 2.7 | 0.14 | 452 | -23 | -38, -7.9 | **0.003** | 583 | -5.1 | -20, 10 | 0.5 | 170 | 27 | -4.6, 59 | 0.093 | 591 | -14 | -28, 0.15 | 0.052 | 614 | -4.4 | -20, 11 | 0.6 |
| APOE ε4 (number) | 1,205 | -51 | -92, -11 | **0.013** | 452 | -45 | -109, 18 | 0.2 | 583 | -33 | -91, 25 | 0.3 | 170 | 10 | -101, 122 | 0.9 | 591 | -57 | -112, -1.7 | **0.043** | 614 | -38 | -97, 22 | 0.2 |
| ADAS<sub>13</sub> (points) | 1,205 | -19 | -23, -15 | **<0.001** | 452 | -10 | -19, -1.6 | **0.020** | 583 | -28 | -34, -22 | **<0.001** | 170 | -3.7 | -13, 5.5 | 0.4 | 591 | -20 | -26, -15 | **<0.001** | 614 | -18 | -24, -11 | **<0.001** |
| GDS (points) | 1,205 | -0.20 | -19, 19 | >0.9 | 452 | -26 | -59, 7.4 | 0.13 | 583 | -1.1 | -27, 25 | >0.9 | 170 | 61 | 10, 111 | **0.019** | 591 | -14 | -39, 12 | 0.3 | 614 | 15 | -14, 43 | 0.3 |
| BMI | 1,205 | 9.1 | 4.1, 14 | **<0.001** | 452 | 7.5 | 0.43, 14 | **0.038** | 583 | 14 | 6.3, 21 | **<0.001** | 170 | -6.8 | -23, 9.4 | 0.4 | 591 | 10 | 4.6, 16 | **<0.001** | 614 | 6.7 | -2.3, 16 | 0.14 |
| ICV (mm<sup>3</sup>) | 1,205 | 0.00 | 0.00, 0.00 | **<0.001** | 452 | 0.00 | 0.00, 0.00 | **<0.001** | 583 | 0.00 | 0.00, 0.00 | **<0.001** | 170 | 0.00 | 0.00, 0.00 | **0.006** | 591 | 0.00 | 0.00, 0.00 | **<0.001** | 614 | 0.00 | 0.00, 0.00 | **<0.001** |

ADAS<sub>13</sub>: Alzheimer's Disease Assessment Scale – 13 items, APOE: Apolipoprotein E, BMI: Body Mass Index, CI: Confidence Interval, GDS: Geriatric Depression Scale, HC: Healthy Controls, ICV: Intracranial volume, MCI: Mild Cognitive Impairment, TSH: Thyroid Stimulating Hormone.

**Supplementary Table 4:** Crude and adjusted models of the regression analyses with <u>total Entorhinal cortex volume</u> as the predicted variable and TSH as the predicting variable.

| Characteristics | Total study participants |  |  |  | Healthy controls |  |  |  | Mild cognitive impairment |  |  |  | Dementia |  |  |  | Females |  |  |  | Males |  |  |  |
| --- | --- | --- | --- | --- | --- | --- | --- | --- | --- | --- | --- | --- | --- | --- | --- | --- | --- | --- | --- | --- | --- | --- | --- | --- |
|  | N | Beta | 95% CI | p-value | N | Beta | 95% CI | p-value | N | Beta | 95% CI | p-value | N | Beta | 95% CI | p-value | N | Beta | 95% CI | p-value | N | Beta | 95% CI | p-value |
| **Crude models** |  |  |  |  |  |  |  |  |  |  |  |  |  |  |  |  |  |  |  |  |  |  |  |  |
| TSH (μIU/mL) | 1,348 | 44 | 3.9, 85 | **0.032** | 522 | 42 | -11, 94 | 0.12 | 641 | 80 | 21, 138 | **0.007** | 185 | -79 | -183, 24 | 0.13 | 667 | 15 | -39, 70 | 0.6 | 681 | 57 | -1.1, 116 | 0.054 |
| **Adjusted models** |  |  |  |  |  |  |  |  |  |  |  |  |  |  |  |  |  |  |  |  |  |  |  |  |
| TSH (μIU/mL) | 1,205 | 40 | 5.1, 75 | **0.025** | 452 | 29 | -23, 81 | 0.3 | 583 | 83 | 27, 138 | **0.003** | 170 | -25 | -117, 67 | 0.6 | 591 | 27 | -20, 74 | 0.3 | 614 | 54 | 1.9, 106 | **0.042** |
| Age (years) | 1,205 | -18 | -24, -12 | **<0.001** | 452 | -24 | -34, -14 | **<0.001** | 583 | -13 | -21, -5.1 | **0.001** | 170 | -24 | -39, -10 | **<0.001** | 591 | -12 | -19, -4.5 | **0.002** | 614 | -25 | -34, -17 | **<0.001** |
| Sex | 1,205 |  |  |  | 452 |  |  |  | 583 |  |  |  | 170 |  |  |  | 591 |  |  |  | 614 |  |  |  |
| Female |  | — | — |  |  | — | — |  |  | — | — |  |  | — | — |  |  |  |  |  |  |  |  |  |
| Male |  | 266 | 174, 358 | **<0.001** |  | 190 | 35, 344 | **0.016** |  | 215 | 77, 353 | **0.002** |  | 340 | 80, 600 | **0.011** |  |  |  |  |  |  |  |  |
| Diagnosis | 1,205 |  |  |  |  |  |  |  |  |  |  |  |  |  |  |  | 591 |  |  |  | 614 |  |  |  |
| HC |  | — | — |  |  | — | — |  |  | — | — |  |  | — | — |  |  | — | — |  |  | — | — |  |
| MCI |  | -232 | -329, -136 | **<0.001** |  |  |  |  |  |  |  |  |  |  |  |  |  | -117 | -249, 14 | 0.080 |  | -327 | -471, -183 | **<0.001** |
| Dementia |  | -508 | -693, -322 | **<0.001** |  |  |  |  |  |  |  |  |  |  |  |  |  | -379 | -650, -109 | **0.006** |  | -640 | -898, -382 | **<0.001** |
| Racial profile | 1,205 |  |  |  | 452 |  |  |  | 583 |  |  |  | 170 |  |  |  | 591 |  |  |  | 614 |  |  |  |
| White |  | — | — |  |  | — | — |  |  | — | — |  |  | — | — |  |  | — | — |  |  | — | — |  |
| Black |  | -113 | -291, 65 | 0.2 |  | -224 | -450, 2.2 | 0.052 |  | 97 | -205, 400 | 0.5 |  | -283 | -948, 382 | 0.4 |  | -100 | -304, 103 | 0.3 |  | -113 | -441, 215 | 0.5 |
| Other |  | 11 | -178, 200 | >0.9 |  | 91 | -186, 367 | 0.5 |  | -26 | -327, 276 | 0.9 |  | -11 | -528, 507 | >0.9 |  | 100 | -134, 333 | 0.4 |  | -91 | -403, 220 | 0.6 |
| Education (years) | 1,205 | -4.2 | -20, 11 | 0.6 | 452 | -11 | -35, 14 | 0.4 | 583 | 0.62 | -22, 24 | >0.9 | 170 | -3.4 | -49, 42 | 0.9 | 591 | 3.0 | -18, 24 | 0.8 | 614 | -12 | -35, 11 | 0.3 |
| APOE ε4 (number) | 1,205 | -40 | -101, 21 | 0.2 | 452 | -18 | -123, 87 | 0.7 | 583 | -18 | -106, 70 | 0.7 | 170 | -79 | -240, 82 | 0.3 | 591 | -44 | -126, 39 | 0.3 | 614 | -42 | -131, 48 | 0.4 |
| ADAS<sub>13</sub> (points) | 1,205 | -26 | -32, -20 | **<0.001** | 452 | 0.20 | -14, 15 | >0.9 | 583 | -33 | -42, -24 | **<0.001** | 170 | -30 | -43, -17 | **<0.001** | 591 | -31 | -39, -22 | **<0.001** | 614 | -22 | -31, -12 | **<0.001** |
| GDS (points) | 1,205 | -2.5 | -31, 26 | 0.9 | 452 | -31 | -85, 24 | 0.3 | 583 | 4.9 | -34, 44 | 0.8 | 170 | 21 | -52, 94 | 0.6 | 591 | -42 | -80, -3.5 | **0.033** | 614 | 34 | -9.4, 76 | 0.13 |
| BMI | 1,205 | 4.0 | -3.7, 12 | 0.3 | 452 | 6.9 | -4.7, 18 | 0.2 | 583 | -0.73 | -12, 11 | >0.9 | 170 | 2.8 | -21, 26 | 0.8 | 591 | 5.2 | -3.7, 14 | 0.2 | 614 | 1.9 | -12, 15 | 0.8 |
| ICV (mm<sup>3</sup>) | 1,205 | 0.00 | 0.00, 0.00 | **<0.001** | 452 | 0.00 | 0.00, 0.00 | **<0.001** | 583 | 0.00 | 0.00, 0.00 | **<0.001** | 170 | 0.00 | 0.00, 0.00 | **0.001** | 591 | 0.00 | 0.00, 0.00 | **<0.001** | 614 | 0.00 | 0.00, 0.00 | **<0.001** |

ADAS<sub>13</sub>: Alzheimer's Disease Assessment Scale – 13 items, APOE: Apolipoprotein E, BMI: Body Mass Index, CI: Confidence Interval, GDS: Geriatric Depression Scale, HC: Healthy Controls, ICV: Intracranial volume, MCI: Mild Cognitive Impairment, TSH: Thyroid Stimulating Hormone.

**Supplementary Table 5:** Crude and adjusted models of the regression analyses with <u>left Entorhinal cortex volume</u> as the predicted variable and TSH as the predicting variable.

| Characteristics | Total study participants |  |  |  | Healthy controls |  |  |  | Mild cognitive impairment |  |  |  | Dementia |  |  |  | Females |  |  |  | Males |  |  |  |
| --- | --- | --- | --- | --- | --- | --- | --- | --- | --- | --- | --- | --- | --- | --- | --- | --- | --- | --- | --- | --- | --- | --- | --- | --- |
|  | N | Beta | 95% CI | p-value | N | Beta | 95% CI | p-value | N | Beta | 95% CI | p-value | N | Beta | 95% CI | p-value | N | Beta | 95% CI | p-value | N | Beta | 95% CI | p-value |
| **Crude models** |  |  |  |  |  |  |  |  |  |  |  |  |  |  |  |  |  |  |  |  |  |  |  |  |
| TSH (μIU/mL) | 1,348 | 29 | 6.7, 51 | **0.011** | 522 | 16 | -14, 45 | 0.3 | 641 | 57 | 25, 89 | **<0.001** | 185 | -28 | -89, 33 | 0.4 | 667 | 13 | -16, 42 | 0.4 | 681 | 36 | 3.0, 68 | **0.032** |
| **Adjusted models** |  |  |  |  |  |  |  |  |  |  |  |  |  |  |  |  |  |  |  |  |  |  |  |  |
| TSH (μIU/mL) | 1,205 | 28 | 8.5, 48 | **0.005** | 452 | 8.5 | -20, 37 | 0.6 | 583 | 60 | 30, 91 | **<0.001** | 170 | 4.0 | -54, 62 | 0.9 | 591 | 21 | -4.4, 46 | 0.11 | 614 | 36 | 6.0, 67 | **0.019** |
| Age (years) | 1,205 | -9.4 | -13, -6.2 | **<0.001** | 452 | -11 | -16, -5.9 | **<0.001** | 583 | -6.7 | -11, -2.2 | **0.003** | 170 | -16 | -25, -7.4 | **<0.001** | 591 | -6.1 | -10, -2.2 | **0.002** | 614 | -13 | -18, -8.1 | **<0.001** |
| Sex | 1,205 |  |  |  | 452 |  |  |  | 583 |  |  |  | 170 |  |  |  | 591 |  |  |  | 614 |  |  |  |
| Female |  | — | — |  |  | — | — |  |  | — | — |  |  | — | — |  |  |  |  |  |  |  |  |  |
| Male |  | 149 | 97, 200 | **<0.001** |  | 109 | 24, 194 | **0.012** |  | 117 | 42, 193 | **0.002** |  | 197 | 33, 360 | **0.019** |  |  |  |  |  |  |  |  |
| Diagnosis | 1,205 |  |  |  |  |  |  |  |  |  |  |  |  |  |  |  | 591 |  |  |  | 614 |  |  |  |
| CN |  | — | — |  |  |  |  |  |  |  |  |  |  |  |  |  |  | — | — |  |  | — | — |  |
| MCI |  | -87 | -142, -33 | **0.002** |  |  |  |  |  |  |  |  |  |  |  |  |  | -27 | -98, 44 | 0.5 |  | -141 | -225, -58 | **<0.001** |
| Dementia |  | -262 | -367, -158 | **<0.001** |  |  |  |  |  |  |  |  |  |  |  |  |  | -185 | -331, -40 | **0.013** |  | -337 | -488, -187 | **<0.001** |
| Racial profile | 1,205 |  |  |  | 452 |  |  |  | 583 |  |  |  | 170 |  |  |  | 591 |  |  |  | 614 |  |  |  |
| White |  | — | — |  |  | — | — |  |  | — | — |  |  | — | — |  |  | — | — |  |  | — | — |  |
| Black |  | -132 | -232, -32 | **0.010** |  | -184 | -308, -60 | **0.004** |  | -16 | -182, 149 | 0.8 |  | -280 | -698, 138 | 0.2 |  | -127 | -237, -18 | **0.023** |  | -118 | -309, 73 | 0.2 |
| Other |  | -20 | -127, 86 | 0.7 |  | -71 | -223, 81 | 0.4 |  | 51 | -115, 216 | 0.5 |  | -25 | -350, 300 | 0.9 |  | 16 | -110, 142 | 0.8 |  | -64 | -245, 118 | 0.5 |
| Education (years) | 1,205 | -2.4 | -11, 6.5 | 0.6 | 452 | -8.1 | -22, 5.4 | 0.2 | 583 | 3.7 | -8.9, 16 | 0.6 | 170 | -9.3 | -38, 19 | 0.5 | 591 | -4.4 | -16, 7.1 | 0.4 | 614 | -1.0 | -14, 12 | 0.9 |
| APOE ε4 (number) | 1,205 | -25 | -60, 9.0 | 0.15 | 452 | -14 | -72, 43 | 0.6 | 583 | -18 | -67, 30 | 0.5 | 170 | -42 | -143, 59 | 0.4 | 591 | -20 | -65, 25 | 0.4 | 614 | -28 | -81, 24 | 0.3 |
| ADAS<sub>13</sub> (points) | 1,205 | -13 | -17, -9.5 | **<0.001** | 452 | 1.6 | -6.2, 9.5 | 0.7 | 583 | -17 | -22, -12 | **<0.001** | 170 | -16 | -24, -7.8 | **<0.001** | 591 | -16 | -21, -12 | **<0.001** | 614 | -10 | -16, -4.8 | **<0.001** |
| GDS (points) | 1,205 | -4.3 | -20, 12 | 0.6 | 452 | -14 | -44, 15 | 0.3 | 583 | -0.05 | -22, 21 | >0.9 | 170 | -0.75 | -46, 45 | >0.9 | 591 | -18 | -39, 2.6 | 0.087 | 614 | 7.5 | -17, 33 | 0.6 |
| BMI | 1,205 | 1.3 | -3.0, 5.6 | 0.6 | 452 | 2.1 | -4.3, 8.5 | 0.5 | 583 | -2.0 | -8.3, 4.3 | 0.5 | 170 | 2.8 | -12, 17 | 0.7 | 591 | 0.71 | -4.1, 5.5 | 0.8 | 614 | 1.8 | -6.1, 9.7 | 0.7 |
| ICV (mm<sup>3</sup>) | 1,205 | 0.00 | 0.00, 0.00 | **<0.001** | 452 | 0.00 | 0.00, 0.00 | **<0.001** | 583 | 0.00 | 0.00, 0.00 | **<0.001** | 170 | 0.00 | 0.00, 0.00 | **0.022** | 591 | 0.00 | 0.00, 0.00 | **<0.001** | 614 | 0.00 | 0.00, 0.00 | **<0.001** |

ADAS<sub>13</sub>: Alzheimer's Disease Assessment Scale – 13 items, APOE: Apolipoprotein E, BMI: Body Mass Index, CI: Confidence Interval, GDS: Geriatric Depression Scale, HC: Healthy Controls, ICV: Intracranial volume, MCI: Mild Cognitive Impairment, TSH: Thyroid Stimulating Hormone.

**Supplementary Table 6:**
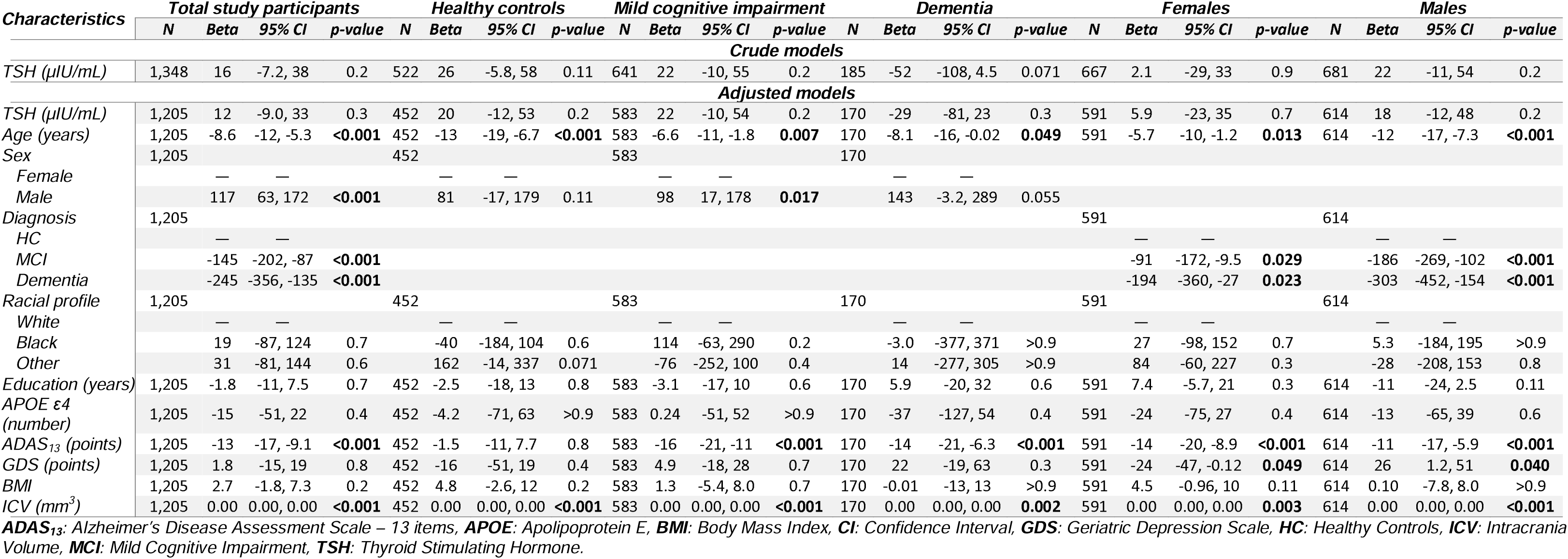
Crude and adjusted models of the regression analyses with <u>right Entorhinal cortex volume</u> as the predicted variable and TSH as the predicting variable.

**Supplementary Table 7:** Crude and adjusted models of the regression analyses with <u>total Amygdala volume</u> as the predicted variable and TSH as the predicting variable.

| Characteristics | Total study participants |  |  |  | Healthy controls |  |  |  | Mild cognitive impairment |  |  |  | Dementia |  |  |  | Females |  |  |  | Males |  |  |  |
| --- | --- | --- | --- | --- | --- | --- | --- | --- | --- | --- | --- | --- | --- | --- | --- | --- | --- | --- | --- | --- | --- | --- | --- | --- |
|  | N | Beta | 95% CI | p-value | N | Beta | 95% CI | p-value | N | Beta | 95% CI | p-value | N | Beta | 95% CI | p-value | N | Beta | 95% CI | p-value | N | Beta | 95% CI | p-value |
| **Crude models** |  |  |  |  |  |  |  |  |  |  |  |  |  |  |  |  |  |  |  |  |  |  |  |  |
| TSH (µIU/mL) | 1,348 | 18 | -9.2, 46 | 0.2 | 522 | 37 | 1.6, 73 | **0.041** | 641 | 23 | -17, 63 | 0.3 | 185 | -67 | -128, -6.4 | **0.030** | 667 | -8.7 | -46, 29 | 0.7 | 681 | 31 | -6.8, 69 | 0.11 |
| **Adjusted models** |  |  |  |  |  |  |  |  |  |  |  |  |  |  |  |  |  |  |  |  |  |  |  |  |
| TSH (µIU/mL) | 1,205 | 16 | -5.2, 37 | 0.14 | 452 | 25 | -7.2, 56 | 0.13 | 583 | 30 | -3.0, 64 | 0.075 | 170 | -43 | -94, 6.9 | 0.090 | 591 | 6.0 | -22, 34 | 0.7 | 614 | 28 | -3.4, 60 | 0.080 |
| Age (years) | 1,205 | -20 | -24, -17 | **<0.001** | 452 | -22 | -28, -16 | **<0.001** | 583 | -22 | -27, -17 | **<0.001** | 170 | -10 | -18, -2.6 | **0.009** | 591 | -18 | -22, -13 | **<0.001** | 614 | -23 | -28, -18 | **<0.001** |
| Sex | 1,205 |  |  |  | 452 |  |  |  | 583 |  |  |  | 170 |  |  |  | 591 |  |  |  | 614 |  |  |  |
| Female |  | — | — |  |  | — | — |  |  | — | — |  |  | — | — |  |  | — | — |  |  | — | — |  |
| Male |  | 235 | 179, 290 | **<0.001** |  | 80 | -15, 175 | 0.10 |  | 261 | 177, 344 | **<0.001** |  | 268 | 126, 410 | **<0.001** |  |  |  |  |  |  |  |  |
| Diagnosis | 1,205 |  |  |  |  |  |  |  |  |  |  |  |  |  |  |  | 591 |  |  |  | 614 |  |  |  |
| HC |  | — | — |  |  | — | — |  |  | — | — |  |  | — | — |  |  | — | — |  |  | — | — |  |
| MCI |  | -162 | -221, -103 | **<0.001** |  |  |  |  |  |  |  |  |  |  |  |  |  | -179 | -259, -100 | **<0.001** |  | -153 | -241, -65 | **<0.001** |
| Dementia |  | -342 | -455, -229 | **<0.001** |  |  |  |  |  |  |  |  |  |  |  |  |  | -299 | -463, -135 | **<0.001** |  | -389 | -547, -231 | **<0.001** |
| Racial profile | 1,205 |  |  |  | 452 |  |  |  | 583 |  |  |  | 170 |  |  |  | 591 |  |  |  | 614 |  |  |  |
| White |  | — | — |  |  | — | — |  |  | — | — |  |  | — | — |  |  | — | — |  |  | — | — |  |
| Black |  | -47 | -155, 60 | 0.4 |  | -63 | -203, 76 | 0.4 |  | 92 | -91, 276 | 0.3 |  | -297 | -661, 66 | 0.11 |  | -25 | -149, 98 | 0.7 |  | -73 | -272, 127 | 0.5 |
| Other |  | -7.3 | -122, 107 | >0.9 |  | 87 | -84, 259 | 0.3 |  | -54 | -236, 129 | 0.6 |  | -21 | -304, 261 | 0.9 |  | 54 | -88, 196 | 0.5 |  | -92 | -282, 98 | 0.3 |
| Education (years) | 1,205 | 0.28 | -9.3, 9.8 | >0.9 | 452 | -3.5 | -19, 12 | 0.7 | 583 | -1.4 | -15, 13 | 0.8 | 170 | 11 | -14, 36 | 0.4 | 591 | -4.5 | -18, 8.5 | 0.5 | 614 | 3.1 | -11, 17 | 0.7 |
| APOE ε4 (number) | 1,205 | -34 | -71, 3.1 | 0.073 | 452 | -14 | -79, 51 | 0.7 | 583 | -9.3 | -63, 44 | 0.7 | 170 | -45 | -133, 43 | 0.3 | 591 | 4.9 | -45, 55 | 0.8 | 614 | -64 | -119, -10 | **0.020** |
| ADAS<sub>13</sub> (points) | 1,205 | -17 | -21, -14 | **<0.001** | 452 | -5.4 | -14, 3.5 | 0.2 | 583 | -23 | -29, -18 | **<0.001** | 170 | -11 | -18, -3.5 | **0.004** | 591 | -20 | -25, -15 | **<0.001** | 614 | -14 | -20, -8.2 | **<0.001** |
| GDS (points) | 1,205 | -10 | -27, 7.2 | 0.3 | 452 | -28 | -62, 5.7 | 0.10 | 583 | -10 | -34, 14 | 0.4 | 170 | 22 | -18, 62 | 0.3 | 591 | -12 | -35, 12 | 0.3 | 614 | -7.8 | -34, 18 | 0.6 |
| Anxiety (binary) | 1,205 | 19 | -53, 92 | 0.6 | 452 | 26 | -165, 217 | 0.8 | 583 | 18 | -81, 117 | 0.7 | 170 | 4.8 | -129, 139 | >0.9 | 591 | -33 | -138, 71 | 0.5 | 614 | 61 | -41, 164 | 0.2 |
| BMI | 1,205 | 6.9 | 2.2, 11 | **0.004** | 452 | 8.2 | 1.0, 15 | **0.025** | 583 | 5.3 | -1.7, 12 | 0.14 | 170 | 2.8 | -9.9, 16 | 0.7 | 591 | 8.9 | 3.6, 14 | **0.001** | 614 | 2.5 | -5.8, 11 | 0.6 |
| ICV (mm<sup>3</sup>) | 1,205 | 0.00 | 0.00, 0.00 | **<0.001** | 452 | 0.00 | 0.00, 0.00 | **<0.001** | 583 | 0.00 | 0.00, 0.00 | **<0.001** | 170 | 0.00 | 0.00, 0.00 | **<0.001** | 591 | 0.00 | 0.00, 0.00 | **<0.001** | 614 | 0.00 | 0.00, 0.00 | **<0.001** |

**ADAS<sub>13</sub>:** Alzheimer's Disease Assessment Scale – 13 items, **APOE:** Apolipoprotein E, **BMI:** Body Mass Index, **CI:** Confidence Interval, **GDS:** Geriatric Depression Scale, **HC:** Healthy Controls, **ICV:** Intracranial volume, **MCI:** Mild Cognitive Impairment, **TSH:** Thyroid Stimulating Hormone.

**Supplementary Table 8:**
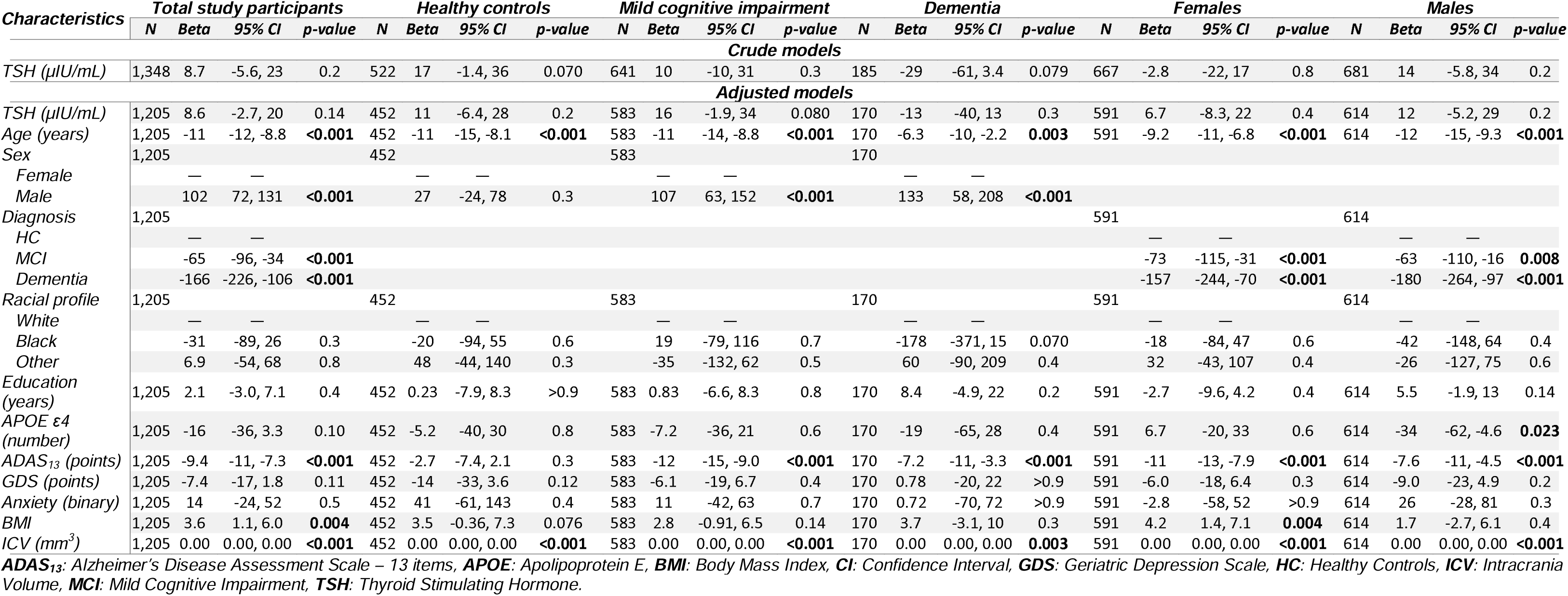
Crude and adjusted models of the regression analyses with <u>left Amygdala volume</u> as the predicted variable and TSH as the predicting variable.

**Supplementary Table 9:** Crude and adjusted models of the regression analyses with <u>right Amygdala volume</u> as the predicted variable and TSH as the predicting variable.

| Characteristics | Total study participants |  |  |  | Healthy controls |  |  |  | Mild cognitive impairment |  |  |  | Dementia |  |  |  | Females |  |  |  | Males |  |  |  |
| --- | --- | --- | --- | --- | --- | --- | --- | --- | --- | --- | --- | --- | --- | --- | --- | --- | --- | --- | --- | --- | --- | --- | --- | --- |
|  | N | Beta | 95% CI | p-value | N | Beta | 95% CI | p-value | N | Beta | 95% CI | p-value | N | Beta | 95% CI | p-value | N | Beta | 95% CI | p-value | N | Beta | 95% CI | p-value |
| **Crude models** |  |  |  |  |  |  |  |  |  |  |  |  |  |  |  |  |  |  |  |  |  |  |  |  |
| TSH (μIU/mL) | 1,348 | 9.5 | -5.0, 24 | 0.2 | 522 | 20 | 0.99, 39 | **0.039** | 641 | 12 | -9.3, 34 | 0.3 | 185 | -39 | -72, -5.2 | **0.024** | 667 | -5.9 | -25, 14 | 0.6 | 681 | 17 | -3.2, 37 | 0.10 |
| **Adjusted models** |  |  |  |  |  |  |  |  |  |  |  |  |  |  |  |  |  |  |  |  |  |  |  |  |
| TSH (μIU/mL) | 1,205 | 7.4 | -4.4, 19 | 0.2 | 452 | 14 | -3.4, 31 | 0.12 | 583 | 14 | -4.2, 33 | 0.13 | 170 | -30 | -59, -0.56 | **0.046** | 591 | -0.68 | -17, 15 | >0.9 | 614 | 17 | -0.95, 34 | 0.064 |
| Age (years) | 1,205 | -9.8 | -12, -7.9 | **<0.001** | 452 | -11 | -14, -7.4 | **<0.001** | 583 | -11 | -13, -8.0 | **<0.001** | 170 | -4.1 | -8.7, 0.46 | 0.078 | 591 | -8.7 | -11, -6.2 | **<0.001** | 614 | -11 | -14, -8.4 | **<0.001** |
| Sex | 1,205 |  |  |  | 452 |  |  |  | 583 |  |  |  | 170 |  |  |  | 591 |  |  |  | 614 |  |  |  |
| Female |  | — | — |  |  | — | — |  |  | — | — |  |  | — | — |  |  |  |  |  |  |  |  |  |
| Male |  | 133 | 102, 164 | **<0.001** |  | 52 | 0.24, 104 | **0.049** |  | 153 | 106, 200 | **<0.001** |  | 135 | 53, 218 | **0.002** |  |  |  |  |  |  |  |  |
| Diagnosis | 1,205 |  |  |  |  |  |  |  |  |  |  |  |  |  |  |  | 591 |  |  |  | 614 |  |  |  |
| HC |  | — | — |  |  | — | — |  |  | — | — |  |  | — | — |  |  | — | — |  |  | — | — |  |
| MCI |  | -97 | -129, -64 | **<0.001** |  |  |  |  |  |  |  |  |  |  |  |  |  | -106 | -151, -61 | **<0.001** |  | -90 | -139, -41 | **<0.001** |
| Dementia |  | -176 | -239, -113 | **<0.001** |  |  |  |  |  |  |  |  |  |  |  |  |  | -142 | -234, -50 | **0.003** |  | -208 | -296, -121 | **<0.001** |
| Racial profile | 1,205 |  |  |  | 452 |  |  |  | 583 |  |  |  | 170 |  |  |  | 591 |  |  |  | 614 |  |  |  |
| White |  | — | — |  |  | — | — |  |  | — | — |  |  | — | — |  |  | — | — |  |  | — | — |  |
| Black |  | -16 | -76, 44 | 0.6 |  | -44 | -120, 33 | 0.3 |  | 74 | -29, 176 | 0.2 |  | -119 | -332, 93 | 0.3 |  | -6.9 | -76, 62 | 0.8 |  | -30 | -141, 81 | 0.6 |
| Other |  | -14 | -78, 50 | 0.7 |  | 39 | -55, 133 | 0.4 |  | -19 | -121, 83 | 0.7 |  | -81 | -246, 84 | 0.3 |  | 22 | -58, 101 | 0.6 |  | -66 | -172, 40 | 0.2 |
| Education (years) | 1,205 | -1.8 | -7.1, 3.5 | 0.5 | 452 | -3.7 | -12, 4.6 | 0.4 | 583 | -2.2 | -10, 5.6 | 0.6 | 170 | 2.6 | -12, 17 | 0.7 | 591 | -1.8 | -9.1, 5.5 | 0.6 | 614 | -2.5 | -10, 5.3 | 0.5 |
| APOE ε4 (number) | 1,205 | -18 | -38, 3.1 | 0.10 | 452 | -8.6 | -44, 27 | 0.6 | 583 | -2.1 | -32, 28 | 0.9 | 170 | -26 | -77, 25 | 0.3 | 591 | -1.8 | -30, 26 | 0.9 | 614 | -31 | -61, -0.68 | **0.045** |
| ADAS<sub>13</sub> (points) | 1,205 | -8.1 | -10, -5.9 | **<0.001** | 452 | -2.7 | -7.6, 2.2 | 0.3 | 583 | -11 | -14, -8.3 | **<0.001** | 170 | -3.6 | -7.9, 0.61 | 0.093 | 591 | -9.3 | -12, -6.3 | **<0.001** | 614 | -6.4 | -9.6, -3.2 | **<0.001** |
| GDS (points) | 1,205 | -2.7 | -12, 6.9 | 0.6 | 452 | -13 | -32, 4.9 | 0.2 | 583 | -3.9 | -17, 9.5 | 0.6 | 170 | 21 | -2.2, 44 | 0.075 | 591 | -5.7 | -19, 7.4 | 0.4 | 614 | 1.2 | -13, 16 | 0.9 |
| Anxiety (binary) | 1,205 | 5.5 | -35, 46 | 0.8 | 452 | -15 | -120, 89 | 0.8 | 583 | 6.8 | -48, 62 | 0.8 | 170 | 4.1 | -74, 82 | >0.9 | 591 | -30 | -89, 28 | 0.3 | 614 | 35 | -22, 92 | 0.2 |
| BMI | 1,205 | 3.3 | 0.69, 5.9 | **0.013** | 452 | 4.7 | 0.78, 8.6 | **0.019** | 583 | 2.5 | -1.4, 6.4 | 0.2 | 170 | -0.83 | -8.3, 6.6 | 0.8 | 591 | 4.7 | 1.7, 7.7 | **0.002** | 614 | 0.77 | -3.8, 5.4 | 0.7 |
| ICV (mm<sup>3</sup>) | 1,205 | 0.00 | 0.00, 0.00 | **<0.001** | 452 | 0.00 | 0.00, 0.00 | **<0.001** | 583 | 0.00 | 0.00, 0.00 | **<0.001** | 170 | 0.00 | 0.00, 0.00 | **0.002** | 591 | 0.00 | 0.00, 0.00 | **<0.001** | 614 | 0.00 | 0.00, 0.00 | **<0.001** |

ADAS<sub>13</sub>: Alzheimer's Disease Assessment Scale – 13 items, APOE: Apolipoprotein E, BMI: Body Mass Index, CI: Confidence Interval, GDS: Geriatric Depression Scale, HC: Healthy Controls, ICV: Intracranial volume, MCI: Mild Cognitive Impairment, TSH: Thyroid Stimulating Hormone.

